# Evidence-Based Interventions Across the Life Course for Healthy Aging: A Scoping Review made in Colombia

**DOI:** 10.1101/2025.08.18.25333703

**Authors:** Lina María González B., Luis Eduardo Mojica, Shannon Riveros, Valentina Vanegas Zamora, Anamaría Gómez, María del Pilar Otero, Felipe Chaves, María Alejandra Jaimes, Valery López, Lina Ramírez, Valeria Sánchez

## Abstract

**Background:** Population aging is a major demographic challenge, particularly in low- and middle-income countries, where the burden of chronic and neurocognitive conditions is projected to grow sharply.

**Objectives:** This scoping review aimed to synthesize scientific evidence on interventions that promote healthy aging across the life course. It was conducted following the Joanna Briggs Institute PCC framework.

**Methods:** A systematic search was performed across PubMed, Scielo, EMBASE, PsycINFO, Cochrane, and LiLACS, without language or date restrictions.

**Results:** From 5,808 records screened, 219 studies met the inclusion criteria, comprising systematic reviews, meta-analyses, clinical trials, quasi-experimental, and observational studies. Interventions were classified by thematic domains, including physical and mental health, social participation, education, technology, nutrition, public policy, and cultural aspects.

**Conclusions:** Most studies focused on adult and older populations, with physical and mental health representing the predominant areas addressed, while other dimensions were less represented. Significant gaps remain regarding racially diverse populations, advanced-aged older adults, genetic stratification, and life-course perspectives, underscoring the need for diversified research designs to guide equitable and comprehensive healthy ageing strategies.

## 1. Introduction

Population aging is one of the most important demographic challenges of the 21st century. According to the World Health Organization (WHO), by 2050 people over 60 years will represent 22% of the global population (1). In Latin America and the Caribbean, demographic changes have led to a rapid reversal of the population pyramid, with projections indicating that by 2060, low- and middle-income countries will bear the highest burden of neurocognitive disorders (2). Although the proportion of life spent in good health has remained constant, additional years are often associated with declining health status (1), underscoring the need for substantial adaptations in health, economic, and social systems to ensure the integration and care of older adults (1).

The concept of aging has evolved considerably. Rowe and Kahn (1987) distinguished between “normal” and “successful” aging, moving beyond chronological age to highlight diverse health outcomes (3). Later, Baltes conceptualized aging as a dynamic and adaptive process, while the WHO frameworks on Active and Healthy Aging incorporated environmental and social dimensions (4). These perspectives illustrate a progressive shift toward a multidimensional understanding of aging influenced by societal changes (5).

Healthy aging is currently defined as a process that promotes and maintains the physical, mental, and social capacities of older adults, enabling independence and well-being in later life (6). This framework emphasizes functional capacity as an interaction between intrinsic factors—physical and mental health—and the environmental conditions that support adaptation (7).

In Colombia, population aging is an increasing concern. Research conducted in 2023 by the Saldarriaga Concha Foundation identified the country as being in a moderately advanced stage of aging, driven by declining birth and fertility rates alongside rising life expectancy (8). This trend presents significant challenges for social security systems and care models. Care for older adults continues to fall disproportionately on women, creating a substantial caregiver burden. Moreover, there are gaps in the enforcement of policies aimed at ensuring quality of life and healthy aging, highlighting the need to strengthen their practical implementation (8).

The 2015 SABE survey found that most Colombians over 60 lived in urban areas, had an average of 5.5 years of education, and lacked pension income or stable financial support. Nearly half expressed a negative perception of aging and reported hypertension and depressive symptoms as the most prevalent chronic conditions, followed by visual and hearing impairments that limited functionality (9). By 2023, most of the older population had attained only primary education (10). These findings reveal social and health vulnerabilities that reinforce the importance of understanding the factors promoting healthy aging.

Despite advances in identifying predictors such as physical activity, self-care, family and community networks, and health system support, there remains a need to synthesize evidence on effective interventions, including how life experiences influence adaptation to aging (11,12,13). To address this, the present study conducts a scoping review of the existing literature, with a specific emphasis on low- and middle-income countries, where evidence remains fragmented. The research question guiding this review is: What evidence-based interventions have been shown to promote healthy aging across the life course in low- and middle-income countries? A scoping review was chosen as the most appropriate design, as it allows for a comprehensive mapping of existing literature, identification of knowledge gaps, and synthesis of a wide range of study types.

## 2. Methods

The aim of this study was to address this gap through a systematic literature review to identify actions, interventions, and practices supported by scientific evidence that promote healthy aging. The study protocol was submitted to the Internal Committee of the Department of the Pontificia Universidad Javeriana in January 2025 and received ethical approval. The review was designed using the PCC framework (Population, Concept, and Context) proposed by the Joanna Briggs Institute, which enables a structured and precise search (9). The Population considered was the lifespan; the Concept included actions, interventions, and evidence-based practices aimed at promoting healthy aging; and the Context encompassed both urban and rural settings

### 2.1.1 Search Strategy

A systematic search was conducted in January 2025 across multiple databases, including PubMed, Scielo, EMBASE, PsycINFO, the Cochrane Central Register of Controlled Trials (via Ovid), and LILACS.The reference lists of all included sources were screened to identify additional studies. Articles published in Spanish and English were considered, as the research team was fluent in both languages, and no publication date limits were applied.

The initial search was restricted to PubMed to identify key studies and develop a structured search strategy. MeSH and EMTREE terms, along with free-text keywords, were selected from the controlled vocabularies of PubMed, Scielo, EMBASE, and PsycNet. Additional search terms were identified using tools such as PubReMiner and MeSH, as well as from abstracts of relevant articles already known to the authors. Search equations and corresponding results are presented in Table 1.

**Table 1.** Search equation strategies.

| No. | Databas<br>e | Search Strategy | Restrictions | Number<br>of Results |
| --- | --- | --- | --- | --- |
| 1 | Pubmed | ((("Healthy Aging"[MeSH] OR "Aging"[MeSH] OR "Longevity"[MeSH] OR "healthy aging"[Title/Abstract] OR "aging"[Title/Abstract] OR "ageing"[Title/Abstract] OR "envejecimiento saludable"[Title/Abstract] OR "envejecimiento"[Title/Abstract])) AND ("Health Promotion"[MeSH] OR "Life Style"[MeSH] OR "Social Determinants of Health"[MeSH] OR "Psychosocial Intervention"[MeSH] OR "Social Work"[MeSH] OR "psychosocial intervention"[Title/Abstract] OR "psychological intervention"[Title/Abstract] OR "intervenciones psicosociales"[Title/Abstract] OR "intervenciones psicológicas"[Title/Abstract] OR "intervenciones"[Title/Abstract] OR "estilo de vida"[Title/Abstract])) | Free full text, Clinical trial, meta-analysis, randomized controlled trial, review, systematic review. English and Spanish | 1238 |
| 1 | Scielo | (ti:(("envejecimiento saludable") OR ("envejecimiento") AND ("intervenciones") OR ("estilo de vida")))) | Spanish and English | 232 |
| 2 | Scielo | ((("Healthy Aging"[MeSH] OR "Aging"[MeSH] OR "Longevity"[MeSH] OR "healthy aging"[Title/Abstract] OR "aging"[Title/Abstract] OR "ageing"[Title/Abstract] OR "envejecimiento saludable"[Title/Abstract] OR "envejecimiento"[Title/Abstract])) AND ("Health Promotion"[MeSH] OR "Life Style"[MeSH] OR "Social Determinants of Health"[MeSH] OR "Psychosocial Intervention"[MeSH] OR "Social Work"[MeSH] OR "psychosocial intervention"[Title/Abstract] OR "psychological intervention"[Title/Abstract] OR "intervenciones psicosociales"[Title/Abstract] OR "intervenciones psicológicas"[Title/Abstract] OR "intervenciones"[Title/Abstract] OR "estilo de vida"[Title/Abstract])) | — | 0 |
| 1 | Embase | ('healthy aging'/exp OR 'healthy aging' OR 'aging'/exp OR 'aging' OR 'longevity'/exp OR 'longevity' OR 'healthy aging':ti,ab OR 'aging':ti,ab OR 'ageing':ti,ab OR 'envejecimiento saludable':ti,ab OR 'envejecimiento':ti,ab) AND ('health promotion'/exp OR 'health promotion' OR 'life style'/exp OR 'life style' OR 'social determinants of health'/exp OR 'social determinants of health' OR 'psychosocial intervention'/exp OR 'psychosocial intervention' OR 'social work'/exp OR 'social work' OR 'psychosocial intervention':ti,ab OR<br>'psychological intervention':ti,ab OR<br>'intervenciones psicosociales':ti,ab OR<br>'intervenciones psicológicas':ti,ab OR<br>'intervenciones':ti,ab OR 'estilo de vida':ti,ab)<br>AND ([cochrane review]/lim OR [systematic<br>review]/lim OR [meta analysis]/lim OR<br>[controlled clinical trial]/lim OR [randomized<br>controlled trial]/lim) AND ([article]/lim OR<br>[review]/lim) AND ([english]/lim OR<br>[spanish]/lim) AND [humans]/lim AND<br>[abstracts]/lim AND ([embase]/lim OR<br>[medline]/lim OR [pubmed-not-<br>medline]/lim) | Study type | 1863 |
| 1 | APA<br>Psycnet | ((TI("healthy aging") OR TI("aging") OR<br>TI("ageing") OR TI("envejecimiento<br>saludable") OR TI("envejecimiento")<br>OR<br>AB("healthy aging") OR AB("aging") OR<br>AB("ageing") OR AB("envejecimiento<br>saludable") OR AB("envejecimiento"))<br>AND<br>(TI("health promotion") OR TI("life style")<br>OR TI("social determinants of health") OR<br>TI("psychosocial intervention") OR<br>TI("social work") OR TI("psychological<br>intervention") OR TI("intervenciones<br>psicosociales") OR TI("intervenciones<br>psicológicas") OR TI("intervenciones") OR<br>TI("estilo de vida")<br>OR<br>AB("health promotion") OR AB("life<br>style") OR AB("social determinants of<br>health") OR AB("psychosocial intervention")) | Peer reviewed<br>– Full | 1072 |
|  |  | OR AB("social work") OR<br>AB("psychological intervention") OR<br>AB("intervenciones psicosociales") OR<br>AB("intervenciones psicológicas") OR<br>AB("intervenciones") OR AB("estilo de vida"))<br>AND<br>(lang: "English" OR lang: "Spanish") |  |  |
| 1 | Cochrane Central Register of Controlled Trials Search Strategy | (("Healthy Aging" OR "Aging" OR "Longevity" OR "healthy aging" OR "aging" OR "ageing" OR "envejecimiento saludable" OR "envejecimiento")<br>AND<br>("Health Promotion" OR "Life Style" OR "Social Determinants of Health" OR "Psychosocial Intervention" OR "Social Work" OR "psychosocial intervention" OR "psychological intervention" OR "intervenciones psicosociales" OR "intervenciones psicológicas" OR "intervenciones" OR "estilo de vida")<br>AND<br>("review" OR "systematic review" OR "randomized controlled trial")) | — | 112 |
| — | LILACS | (("Healthy Aging" OR "Aging" OR "Longevity" OR "healthy aging" OR "aging" OR "ageing" OR "envejecimiento saludable" OR "envejecimiento")<br>AND<br>("Health Promotion" OR "Life Style" OR "Social Determinants of Health" OR "Psychosocial Intervention" OR "Social Work" OR "psychosocial intervention" OR "psychological intervention" OR | Full text in English or Spanish" | 1510 |
|  |  | "intervenciones psicosociales" OR<br>"intervenciones psicológicas" OR<br>"intervenciones" OR "estilo de vida")) |  |  |
| — | ProQuest<br>Search<br>Strategy | ((TI("healthy aging") OR TI("aging") OR<br>TI("ageing") OR TI("envejecimiento<br>saludable") OR TI("envejecimiento")<br>OR<br>AB("healthy aging") OR AB("aging") OR<br>AB("ageing") OR AB("envejecimiento<br>saludable") OR AB("envejecimiento"))<br>AND<br>(TI("health promotion") OR TI("life style")<br>OR TI("social determinants of health") OR<br>TI("psychosocial intervention") OR<br>TI("social work") OR TI("psychological<br>intervention") OR TI("intervenciones<br>psicosociales") OR TI("intervenciones<br>psicológicas") OR TI("intervenciones") OR<br>TI("estilo de vida")<br>OR<br>AB("health promotion") OR AB("life<br>style") OR AB("social determinants of<br>health") OR AB("psychosocial intervention")<br>OR AB("social work") OR<br>AB("psychological intervention") OR<br>AB("intervenciones psicosociales") OR<br>AB("intervenciones psicológicas") OR<br>AB("intervenciones") OR AB("estilo de<br>vida"))))<br>AND<br>LA(("English" OR "Spanish")) | Peer-reviewed<br>scientific<br>journals, full<br>text in Spanish<br>and English | 13 |

### 2.1.2 Inclusion Criteria

Quantitative studies evaluating interventions to promote healthy aging, conducted in urban and/or rural settings, were included regardless of the country of origin. Eligible interventions targeted all age groups, with particular attention to those involving individuals with non-communicable chronic diseases, especially metabolic conditions such as obesity and diabetes mellitus. Studies were also required to report outcomes related to quality of life, functional capacity, and physical, mental, or social well-being. Only peer-reviewed, full-text articles published in Spanish or English were considered, with no restrictions on publication date.

### 2.1.3 Exclusion Criteria

Research protocols, ongoing studies without conclusive results, and those not subjected to peer review were excluded, as the review focused on interventions reporting measurable and comparable outcomes related to healthy aging. Studies exclusively targeting individuals with cognitive impairment, psychotic disorders, or affective disorders were also excluded, as were opinion pieces, editorials, and unpublished theses. Additionally, studies unavailable in full text or lacking sufficient information for analysis were discarded. Qualitative studies were excluded as well, given that the objective of this review was to synthesize evidence based on quantifiable outcomes—such as quality of life, functional capacity, and physical, mental, or social well-being. This decision enabled a structured comparison and categorization of interventions across studies, facilitating the identification of patterns and gaps in the existing evidence

### 2.1.4 Data Charting, Collection, Summarization

After completing the literature search, all retrieved citations were uploaded into Rayyan QCRI, which facilitated duplicate removal through its automated detection function. A manual verification was also performed to ensure accuracy.

Prior to the formal selection process, a pilot test was conducted to ensure that reviewers shared a consistent understanding of the inclusion criteria. The selection process was subsequently carried out in two stages: titles and abstracts were independently screened by at least two reviewers according to the eligibility criteria, and articles meeting these criteria underwent full-text evaluation. Reasons for exclusion were documented and reported to maintain transparency in the scoping review.

For each included study, summary tables were created to facilitate analysis and comparison. These tables captured bibliographic details (title, authors, year, country), study type, objectives, main findings, conclusions, funding sources, and assessments of risk of bias and conflicts of interest. Methodological quality observations were also recorded. Each intervention was categorized by age group and detailed according to activity type, duration, location, cultural adaptations, and long-term follow-up. Main outcomes focused on quality of life, functional capacity, and indicators of physical, mental, and social well-being. To synthesize the findings, thematic categories were developed, including primary, secondary, and tertiary domains such as physical health, mental health, social participation, nutrition, technology, environment, education, and public policy. This systematization enabled a structured and comprehensive characterization of the available evidence on healthy aging promotion.

Following article selection, artificial intelligence (ChatGPT) was employed solely as an auxiliary tool to facilitate structured data extraction. Its use was limited to organizing and systematizing information, without any influence on the study results or interpretation. To ensure reliability, researchers manually reviewed 10% of the articles, confirming the accuracy and consistency of the AI-assisted extraction. Studies that did not meet the methodological standards established for this review were excluded.

The entire search and selection process, along with study retrieval and inclusion, is presented in a flow diagram following the Preferred Reporting Items for Systematic Reviews and Meta-Analyses for Scoping Reviews (PRISMA-ScR) guidelines (Figure 1). This diagram illustrates each stage of the review, including duplicate removal, full-text screening, additional sources, data extraction, and evidence synthesis, ensuring transparency and methodological rigor

**Figure 1.**
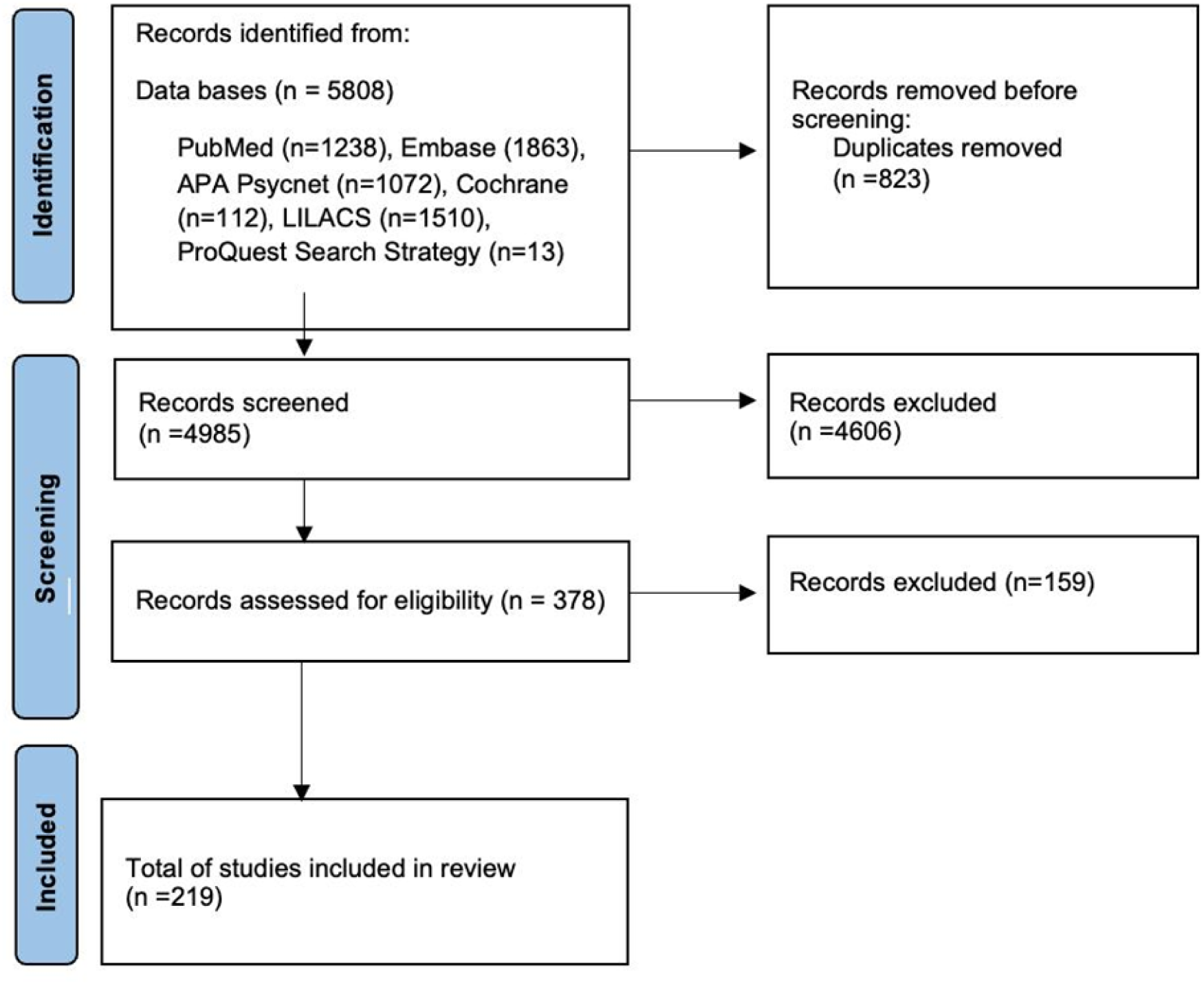
PRISMA flow diagram

## Results

A total of 5,808 records were identified through searches in scientific databases and grey literature. After removing 823 duplicate records, 4,986 titles and abstracts were screened. Of these, 379 full-text articles were assessed for eligibility. Ultimately, 219 studies met the inclusion criteria and were included in the final review. No additional sources were identified through manual searching or reference list screening.

The included studies encompassed various designs, including systematic reviews, meta-analyses, scoping reviews, clinical trials, quasi-experimental studies, and observational research. Together, they examined a broad spectrum of interventions aimed at promoting healthy aging across different stages of the life course. Findings were organized by study design, type of intervention, reported outcomes, limitations, and conclusions. The studies were also grouped into nine thematic categories reflecting the most recurrent areas and recommendations: physical and mental health, social behaviors, education, technology use, public policy and the built environment, religion and spirituality, nutrition, life course perspectives, and other relevant domains. A total of 78 studies focused on physical health and 43 on mental and emotional health, representing the most frequently addressed areas in the reviewed literature. Other domains were less frequently represented, including social networks and participation (n=31), education and development (n=16), technology (n=12), environmental and public policy (n=15), nutrition (n=15), life-course approach (n=6), and religion and culture (n=2). Most studies targeted adult and older adult populations, with limited representation of early or mid-life stages.

Due to the volume of findings and the varying quality of evidence, only the most relevant results are presented in this section.

## Discussion

The dominance of evidence related to physical and mental health reflects a prevailing biomedical orientation in the literature on healthy aging. In contrast, the limited representation of domains such as religion, nutrition, and life-course approaches constrains the development of multidimensional and culturally contextualized models. Moreover, the disproportionate focus on adult and older adult populations reinforces a segmented conceptualization of ageing, often detached from early and mid-life stages where foundational determinants of later-life outcomes are shaped. This age-bounded perspective may hinder the formulation of comprehensive, life-course-informed public health strategies.

### Life-course approach

Reviewed studies that have a life course approach predominantly employ observational designs, including cross-sectional (14,15), longitudinal (16,17,18), and mixed-cohort analyses (19). While the predominance of observational evidence limits causal inference, it offers notable strengths, including large population-based samples, probabilistic recruitment, repeated assessments over time, and the use of validated psychometric instruments. Collectively, these features enhance ecological validity and facilitate the exploration of developmental patterns across the life course.

The findings converge on multiple psychosocial, behavioral, and contextual determinants of well-being and cognitive health in adulthood, though the methodological constraints temper causal interpretation. Reference (14) found that the apparent positive association between age and job satisfaction was largely attributable to job congruence, locus of control, tenure, and salary, supporting the job change hypothesis and suggesting that occupational alignment, rather than chronological age, underpins workplace satisfaction in later life. In (15), the authors identified future-oriented planning as a mediator linking education, conscientiousness, and perceived control to enhanced life satisfaction, particularly in older adults, highlighting the value of proactive coping strategies in sustaining psychological health. The analysis in (19) showed that emotional stability consistently predicts life, work, and social satisfaction across adulthood, with conscientiousness most strongly linked to job satisfaction and extraversion and agreeableness to social satisfaction, highlighting personality traits as stable yet potentially adaptable factors influencing subjective well-being.

Beyond psychosocial factors, (16) describes a longitudinal study in Germany with 100 cognitively healthy participants aged 10–79 years, assessing episodic memory retention 11 months after mnemonic training through age-stratified comparisons and validated measures. The results, showing ongoing improvement in children and context-dependent retention in older adults, highlight the importance of a life-course perspective in understanding cognitive aging. According to (17), an observational study of 150 U.S. adults aged 18–89 years found that daily physical activity was consistently linked to higher life satisfaction across adulthood, while cumulative habitual activity had stronger benefits in midlife and later life, effects partly explained by physical and mental health. These associations, identified through daily longitudinal assessments and multivariate modeling, remained significant even after accounting for daily health fluctuations. Data from (18), involving 8,538 U.S. adults aged 45– 93 years in the REGARDS cohort, showed that retrospectively reported childhood social support predicted better baseline episodic memory in later life, though not verbal fluency or cognitive change over 10 years. These effects, identified through latent trajectory modeling, were partly mediated by higher educational attainment, lower stress, and lower body mass index, highlighting how early social environments can shape cognitive health decades later and reinforcing the importance of a life-course perspective in understanding aging.

Nonetheless, the lack of experimental designs significantly restricts the ability to draw causal conclusions. Cross-sectional studies cannot establish temporal ordering, and even longitudinal analyses remain vulnerable to unmeasured confounding and selection biases. Consequently, while the evidence highlights potentially modifiable pathways to healthy ageing—such as physical activity, personality-linked traits, social support, and cognitive training—its strength is tempered by the inherent methodological limitations in establishing directionality and efficacy

### Physical health

Multicomponent interventions that combine strength, balance, endurance, and flexibility exercises have shown consistent effects on physical function in older adults. Reference (20) reported significant improvements in muscle strength, gait speed, balance, and functional independence among institutionalized older adults, with the greatest effect observed with 170 minutes of exercise per week. Other community-based programs for pre-frail older adults led to significant improvements in the achievement of individual goals, as measured by the Goal Attainment Scaling (GAS) (21). Likewise, authors in (22) confirmed that different types of training (strength, balance, and others), as well as their combination, have a positive impact on muscle strength and balance, although there is no conclusive evidence on the maintenance of these effects. Evidence focused on habitual physical activity rather than structured programs similarly highlights the preventive effects against disability in basic activities of daily living among community-dwelling older adults who are moderately to highly physically active (23).

Regarding specific exercise modalities, comparative analyses of high-intensity interval training (HIIT) and moderate-intensity continuous training (MICT) found significant improvements in cardiorespiratory fitness and metabolic parameters with both, and greater benefits in body composition and cognitive performance with HIIT (24). These findings complement those from a study that evaluated the combination of physical exercise and nutritional supplementation in menopausal women, identifying synergistic improvements in muscle mass, strength, and vasomotor symptoms, especially when strategies such as resistance training or HIIT were employed (25). A review of more than 1,400 studies confirmed that structured exercise was more effective in preventing falls and improving functional capacity, while recreational physical activities (such as dance, tai chi, and yoga) had a greater impact on cognitive and emotional domains (26).

Activities with a cultural or recreational component also show relevant positive effects, as documented through improvements in balance, strength, and flexibility in healthy older adults participating in programs involving traditional dance, Muay Thai, or line dancing (27). Similarly, Olympic combat sports such as karate and judo improved health-related quality of life, particularly physical and emotional aspects (28). Studies on Taekwondo have reported significant cognitive, emotional, and functional benefits, supporting its value as a comprehensive intervention. Similarly, yoga has been associated with improvements in mental well-being, functional balance, and perceived health (29,30). Furthermore, additional sources report that Tai Chi and Qigong have consistent effects on physical functioning, psychological well-being, and fall prevention in older adults, with or without chronic conditions (31).

In workplace settings, programs aimed at workers over 50 years of age showed modest improvements in strength, balance, and flexibility through interventions such as walking, yoga, and resistance band exercises (32). Although the methodological quality was low, the results are consistent with those observed in community and institutional settings. Similar effects were found for the Pilates method on balance, autonomy, and flexibility, albeit with lower methodological standardization (33).

Population-based and biomarker-oriented studies further enrich the evidence. For instance, research from (34) found that higher levels of moderate-to-vigorous physical activity were inversely associated with sarcopenia, whereas sedentary behavior showed less consistent associations. In addition, findings from (35) indicated that physical activity is negatively associated with metabolomic biomarkers of aging such as MetaboAge and MetaboHealth, suggesting its value as a modifiable factor of biological aging. Among the oldest-old (≥80 years), studies have shown that regular exercise, alongside diet and leisure activities, protects against cognitive decline even in advanced stages of life (36). Complementarily, analyses conducted in Latin America showed that physical activity is one of the most consistent predictors of healthy aging, along with social support and education (37).

Finally, integrative approaches combining physical activity with music suggest improvements in cognitive functions such as verbal memory and fluency, although based on heterogeneous studies (38). A Cochrane systematic review also reported that behavioral and educational interventions can reduce sedentary time in older adults, although with modest effects and low-quality evidence (39). While these studies are less robust, they indirectly support the usefulness of promoting active lifestyles as a strategy for healthy aging.

### Mental health

Evidence on interventions for healthy ageing remains dominated by systematic reviews and meta-analyses of randomized controlled trials, but a closer examination of each intervention highlights distinct patterns of outcomes and limitations.

Dance-based programs (40,41) consistently reported improvements in executive function, processing speed, and balance, along with positive effects on mood and quality of life. Benefits were most evident in programs combining structured choreography, music, and social interaction. However, the majority of trials included small, homogeneous samples of healthy, community-dwelling older adults, with intervention periods averaging 8–12 weeks and limited follow-up, which constrains conclusions about long-term maintenance of gains and applicability in individuals with multimorbidity.

Computerized cognitive training and memory strategy instruction (42,43) demonstrated modest but significant improvements in working memory, attention, and self-perceived cognitive abilities. Gains were more consistent in programs with frequent sessions (≥3 times/week) and adaptive difficulty. Nonetheless, reviews identified heterogeneity in training platforms, outcome measures, and control conditions, as well as a lack of clarity regarding optimal training dose and duration to sustain effects over time.

External memory aids and compensatory strategies showed enhanced task performance and adherence to daily activities in structured assessments (44). However, these effects were task-specific, with limited evidence of transfer to broader functional independence. Short intervention durations and suboptimal blinding in many trials further limit confidence in generalizing these findings to real-world contexts.

Psychological and multimodal cognitive interventions reported small to moderate effects on memory, reasoning, and self-efficacy, particularly when interventions integrated group-based activities, social engagement, and problem-solving tasks (45) The diversity of protocols and small number of included studies complicated meta-analytic synthesis, reducing certainty of pooled estimates.

Critical appraisal of these results shows that while interventions yield statistically significant cognitive and psychosocial benefits, their practical impact remains uncertain due to methodological limitations: short follow-up, selective samples, and high control of intervention environments. To enhance applicability and scalability, future research should prioritize pragmatic and hybrid effectiveness–implementation trials that include diverse ageing populations, extend follow-up periods, and evaluate real-world outcomes such as independence, functional capacity, and sustained engagement.

### Social networks and participation

One systematic review synthesized evidence from 14 intervention studies, predominantly involving quasi-experimental and observational designs, with only three employing randomized allocation (46). The studies aimed at supporting healthy transitions into retirement by fostering meaningful social roles among adults aged 55 to 70. The interventions examined were diverse in format but shared a common emphasis on structured social participation through activities like volunteering in schools, mentoring youth, maintaining public spaces, and participating in intergenerational support networks. By assigning explicit responsibilities and socially valued roles, these programs sought to reinforce older adults’ sense of purpose, belonging, and societal contribution. Programs offering financial incentives reported higher participation rates, indicating the relevance of both intrinsic and extrinsic motivators in promoting sustained engagement. However, the interventions varied substantially in structure, intensity, and delivery, limiting the ability to compare effectiveness across settings

The limited number of RCTs in the review restricts the strength of the conclusions that can be drawn about causality between intervention exposure and improvements in well-being indicators such as life satisfaction, cognitive function, and social support. Additionally, the considerable heterogeneity in intervention modalities, theoretical underpinnings, and evaluation tools further complicates cross-study comparability and limits the generalizability of findings. While the review underscores promising outcomes, especially from programs anchored in psychological and sociological theories, the evidence base lacks the methodological rigor necessary to inform standardized policy recommendations. Furthermore, the absence of qualitative data on participants lived experiences constrains the understanding of how participants interpreted and internalized their roles Thus, advancing this field requires an expansion of high-quality mixed-methods research, with greater emphasis on randomized designs, theory-driven frameworks, and longitudinal follow-up to capture long-term effects and contextual adaptability in diverse ageing populations.

### Education and development

Among the included studies, systematic reviews were the predominant methodological design (47,48). A meta-analysis and systematic review examined the effects of non-exercise interventions on quality of life, encompassing a wide range of non-physical strategies such as lifelong learning, leisure activities, art therapy, social support, mindfulness, health education, technology use, occupational workshops, and home visits. The review noted variability in intervention types and durations, along with concerns about risk of bias and sample representativeness (47). A comprehensive review (48) synthesized diverse health promotion strategies across international contexts and highlighted the inconsistent use of validated instruments and limited statistical detail, which constrains the interpretability and comparability of findings. However, the reliance on systematic reviews also introduces inherent limitations, particularly when the included primary studies are heterogeneous or methodologically weak, as was the case in both reviews.

This methodological distribution affects the robustness and generalizability of the current evidence base in several ways. In both examples, the presence of bias, methodological heterogeneity, and inconsistent outcome measures weakens the overall certainty of the evidence. Second, the limited inclusion of randomized controlled trials (RCTs) or well-designed quasi-experimental studies diminishes the causal inferences that can be drawn regarding intervention effectiveness. Third, the overrepresentation of certain populations raises concerns about the external validity of the findings, especially when generalizing to more diverse or underrepresented subgroups of older adults.

Despite these limitations, the systematic review methodology has contributed valuable insights, particularly by identifying promising non-physical interventions (e.g., art therapy, social engagement, and lifelong learning) that may complement traditional physical activity-based approaches. Nonetheless, to enhance the applicability of findings to real-world settings, future research should prioritize methodologically rigorous primary studies with standardized outcomes, diverse populations, and context-specific implementation strategies. This would strengthen the translational potential of evidence into effective, equitable interventions for healthy ageing across varied sociocultural and health system contexts.

### Technology

Evidence on technology-based interventions for healthy ageing is primarily derived from systematic reviews and meta-analyses, with a strong focus on digital tools to support physical activity, cognitive stimulation, and social engagement. Studies identified digital platforms and mobile applications as effective for increasing physical activity in older adults, particularly programs combining goal setting, real-time feedback, and remote coaching (49,50). Pooled analyses showed significant increases in moderate-to-vigorous physical activity and reductions in sedentary behavior, with web-based and app-based interventions demonstrating the greatest impact.

Similarly, an umbrella review (51) and a focused meta-analysis (52), evaluated video game-based cognitive interventions. Findings consistently indicated improvements in attention, processing speed, and working memory, with action and strategy games showing the most robust cognitive gains. However, effects on global cognition and everyday functioning were less consistent, suggesting a need to refine game design to enhance transferability of skills to daily life.

Other systematic reviews (53,54) highlighted the use of assistive and remote monitoring technologies (e.g., wearable sensors, telehealth platforms) to support fall prevention, medication adherence, and chronic disease management. These interventions were associated with improved self-management, reduced hospital visits, and higher perceived independence, particularly when combined with personalized feedback and caregiver integration.

Overall, the evidence suggests that technology-based interventions are feasible, acceptable, and capable of producing measurable benefits in physical activity, cognitive performance, and functional independence among older adults. However, sustained effectiveness is contingent upon digital literacy, usability of devices, ongoing engagement, and access to infrastructure. To enhance real-world applicability, future research should evaluate long-term adherence, equity of access across socioeconomic groups, and integration into community and healthcare systems.

### Nutrition

The included studies in this review exhibit a predominant reliance on randomized controlled trials (RCTs), with five of the six systematic reviews and meta-analyses incorporating exclusively RCTs or prioritizing them in their analyses (55,56,57,58,59). Two of the reviews (59,60) synthesized data from observational or epidemiological studies, particularly to explore long-term associations in population-based settings. The methodological strengths of this distribution lie in the high internal validity of RCTs, which allow for causal inferences regarding the effectiveness of specific interventions, such as caloric restriction, Mediterranean diet, eHealth nutrition tools, or mind-body practices. These approaches demonstrated consistent benefits in domains like global cognition, cardiometabolic health, and anthropometric indicators, suggesting that well-targeted and moderately intensive interventions can contribute meaningfully to healthy ageing. RCTs also typically incorporate standardized protocols and validated outcome measures, reducing the risk of measurement bias and confounding. However, the dominance of RCTs, particularly those with short to moderate durations and limited sample sizes, may constrain external validity and obscure long-term or population-wide effects relevant to healthy ageing. This is particularly evident in the limited generalizability of findings to very old adults, socioeconomically diverse groups, or individuals with multimorbidity.

Conversely, observational studies, (59,60) provide valuable associations over extended periods and in broader age ranges, including centenarians. These designs enhance ecological validity and capture exposures that are difficult to evaluate in controlled environments (e.g., habitual fish intake or lifetime medication use). Nonetheless, their susceptibility to residual confounding, reverse causation, and heterogeneity in exposure assessment and outcome measurement weakens the causal interpretability of findings. Moreover, most observational studies included lacked detailed adjustment for covariates, which may compromise the robustness of reported associations.

The current methodological landscape thus reflects a tension between internal and external validity. While RCTs offer rigorous evidence on specific, modifiable interventions, their findings are often derived from controlled conditions that may not translate seamlessly into heterogeneous ageing populations or complex, multimodal public health strategies. In contrast, observational evidence provides breadth and contextual relevance but is methodologically limited in establishing causality. Consequently, while the evidence base supports modest benefits from select interventions—particularly Mediterranean diet components, tai chi, caloric restriction, and multicomponent eHealth approaches—its overall robustness and generalizability remain constrained. Future research should prioritize long-duration, well-powered RCTs employing integrated interventions and stratified analyses to better inform real-world policies and programs promoting healthy ageing.

## Conclusions and implications

Although a substantial portion of the reviewed literature provides descriptive or qualitative insights without clear guidance on actionable strategies, the strongest body of evidence remains concentrated in systematic reviews and meta-analyses of randomized controlled trials, particularly in domains such as cognitive and psychological training, digital interventions, physical activity, nutrition, and social engagement. These studies establish preliminary efficacy across diverse outcomes, including cognitive function, mood, physical health, and quality of life. However, the current evidence base is limited by its frequent exclusion of key subgroups— such as racially diverse populations, advanced-aged old adults, individuals with multimorbidity, and those across earlier life stages—alongside a near absence of genetic or biological stratification in most analyses. Addressing these gaps will require methodologically diversified research, including pragmatic trials, longitudinal and life-course studies, and designs capable of integrating biological, social, and behavioral determinants. Future investigations must also embrace disaggregated analyses by race, age, and genetic markers to clarify differential effects and mechanisms of benefit. Such efforts are crucial for advancing beyond proof-of-concept efficacy and toward tailored, equitable strategies that reflect the heterogeneity of aging trajectories, ultimately guiding the development of comprehensive, evidence-informed policies and interventions for healthy ageing.

To translate existing evidence into public health action, policymakers should prioritize the implementation of multicomponent interventions that go beyond addressing physical and cognitive health to also include social participation, education, nutrition, and environmental support. These programs must be culturally adapted, accessible across socioeconomic strata, and embedded within community infrastructure to ensure sustainability and equity. It is essential to shift away from a disease-centered model and instead emphasize health promotion, prevention, and the strengthening of community-based strategies that foster autonomy and social inclusion. Moreover, these efforts should not be limited to adults and older adults, but integrated from early life stages through a life-course approach that builds the foundations for healthy aging from childhood onwards. Incorporating older adults and caregivers in the design of these interventions is also crucial to enhance their relevance and adoption. Finally, investing in cross-sectoral collaboration between health, social, and educational systems will be key to fostering healthy aging across the life course.

## Data Availability

All data produced in the present study are available upon reasonable request to the authors

## Credit authorship contribution statement

Conceptualization and methodology were carried out by González L and Mojica L. Data curation was performed by Mojica L, Riveros S, Zamora V, Gómez A, Otero M, Chaves F, Jaimes M, López V, Ramírez L, and Sánchez V. Formal analysis and interpretation were conducted by all authors, who also contributed to the drafting and visualization of the manuscript, as well as its critical revision to ensure the quality and relevance of the intellectual content.

## Acknowledgements

The authors would like to express their sincere gratitude to Pontificia Universidad Javeriana (PUJ) for its continuous institutional support and for providing the academic environment that made this study possible. We are especially thankful to the Mental Health Research Group of the PUJ for their valuable contributions, insightful discussions, and commitment to fostering research on evidence-based interventions for healthy aging. Their support was instrumental throughout the development and execution of this scoping review.

## Declaration of Artificial Intelligence Assistance

Artificial intelligence (ChatGPT, OpenAI) was used exclusively to assist in extracting key data from the included studies after article selection. The tool was not employed to generate original content or interpret findings. To ensure reliability and accuracy, 10% of the extracted data were manually verified by the research team, and all inconsistencies were corrected. Final data interpretation, synthesis, and manuscript preparation were conducted entirely by the authors.

## References

1. World Health Organization. Ageing and health. 2023.

2. Comisión Económica para América Latina y el Caribe (CEPAL). Panorama del envejecimiento: tendencias demográficas en América Latina y el Caribe. 2023.

3. Rowe JW, Kahn RL. Human aging: usual and successful. Science. 1987;237(4811):143–149. doi:10.1126/science.3299702

4. Behr LC, Simm A, Kluttig A, Großkopf A. 60 years of healthy aging: on definitions, biomarkers, scores and challenges. Ageing Res Rev. 2023;88:101934. doi:10.1016/j.arr.2023.101934

5. Peters MDJ, Godfrey CM, Khalil H, McInerney P, Parker D, Soares CB. 2017 guidance for the conduct of JBI scoping reviews. JBI Evidence Implementation. 2017;15(4):121–126.

6. World Health Organization. Decade of healthy ageing. 2023.

7. Castro-Suarez S. Envejecimiento saludable y deterioro cognitivo. Rev Neuropsiquiatr. 2018;81(4):215–216. doi:10.20453/rnp.v81i4.3435

8. Fundación Saldarriaga Concha, Fedesarrollo, PROESA, DANE. Misión Colombia Envejece: una investigación viva. Bogotá: Fundación Saldarriaga Concha; 2023.

9. Ministerio de Salud y Protección Social. SABE Colombia 2015: estudio nacional de salud, bienestar y envejecimiento. Bogotá: Minsalud; 2015.

10. Fundación Saldarriaga Concha, Inclusión SAS. Colombia envejece: las oportunidades de una sociedad longeva. Bogotá: Fundación Saldarriaga Concha; 2025.

11. Dziechciaż M, Filip R. Biological psychological and social determinants of old age: bio-psycho-social aspects of human aging. Ann Agric Environ Med. 2014;21(4):835–838. doi:10.5604/12321966.1129943

12. Fried LP, Tangen CM, Walston J, Newman AB, Hirsch C, Gottdiener J, et al. Frailty in older adults: evidence for a phenotype. J Am Geriatr Soc. 2001;50(3):24. doi:10.1046/j.1532-5415.2002.50324.x

13. Bosch JV. Filming “successful aging“. Gerontologist. 2015;55(1):169–170. doi:10.1093/geront/gnu172

14. White AT, Spector PE. An investigation of age-related factors in the age-job-satisfaction relationship. Psychol Aging. 1987;2(1):261–265.

15. Prenda KM, Lachman ME. Planning for the future: a life management strategy for increasing control and life satisfaction in adulthood. Psychol Aging. 2001;16(2):206–216.

16. Brehmer Y, Li SC, Straube B, Stoll G, von Oertzen T, Müller V, et al. Comparing memory skill maintenance across the life span: preservation in adults, increase in children. Psychol Aging. 2008;23(2):227–238.

17. Maher JP, Pincus AL, Ram N, Conroy DE. Daily physical activity and life satisfaction across adulthood. Dev Psychol. 2015;51(10):1407–1419.

18. Zahodne LB, Sharifian N, Manly JJ, Sumner JA, Crowe M, Wadley VG, et al. Life course biopsychosocial effects of retrospective childhood social support and later-life cognition. Psychol Aging. 2019;34(7):867–883.

19. Olaru G, van Scheppingen MA, Bleidorn W, Denissen JJA. The link between personality, global, and domain-specific satisfaction across the adult lifespan. J Pers Soc Psychol. 2023;125(6):1250–1276. doi:10.1037/pspp0000461

20. Valenzuela PL, Saco-Ledo G, Morales JS, Gallardo-Gómez D, Morales-Palomo F, López-Ortiz S, et al. Effects of physical exercise on physical function in older adults in residential care: a systematic review and network meta-analysis of randomised controlled trials. Lancet Healthy Longev. 2023;4(6):e247–e256.

21. Podolski OS, Whitfield T, Schaaf L, Cornaro C, Köbe T, Koch S, Wirth M. The impact of dance movement interventions on psychological health in older adults without dementia: a systematic review and meta-analysis. Brain Sci. 2023;13(7):981. doi:10.3390/brainsci13070981

22. Adams M, Gordt-Oesterwind K, Bongartz M, Zimmermann S, Seide S, Braun V, Schwenk M. Effects of physical activity interventions on strength, balance and falls in middle-aged adults: a systematic review and meta-analysis. Sports Med Open. 2023;9:61. doi:10.1186/s40798-023-00606-3

23. Tak E, Kuiper R, Chorus A, Hopman-Rock M. Prevention of onset and progression of basic ADL disability by physical activity in community dwelling older adults: a meta-analysis. Ageing Res Rev. 2013;12(4):329–338.

24. Oliveira A, Fidalgo A, Farinatti P, Monteiro W. Effects of high-intensity interval and continuous moderate aerobic training on fitness and health markers of older adults: a systematic review and meta-analysis. Arch Gerontol Geriatr. 2024;124:105451. doi:10.1016/j.archger.2024.105451

25. Sánchez-García JC, López Hernández D, Piqueras-Sola B, Cortés-Martín J, Reinoso-Cobo A, Menor-Rodríguez MJ, Rodríguez-Blanque R. Physical exercise and dietary supplementation in middle-aged and older women: a systematic review. J Clin Med. 2023;12(23):7271. doi:10.3390/jcm12237271

26. Pinheiro MB, Sherrington C, Oliveira JS, Ramsay E, Chamberlain K, Paul SS, et al. Physical activity interventions for older adults: a rapid review. Int J Behav Nutr Phys Act. 2022;19:87. doi:10.1186/s12966-022-01297-2

27. Sooktho S, Lawanprasert S, Jongkulsathain R. A meta-analysis of the effects of dance programs on physical function in healthy older adults. Ann Geriatr Med Res. 2022;26(3):196–207. doi:10.4235/agmr.22.0053

28. Valdés-Badilla P, Herrera-Valenzuela T, Guzmán-Muñoz E, Delgado-Floody P, Núñez-Espinosa C, Monsalves-Álvarez M, et al. Effects of Olympic combat sports on health-related quality of life in middle-aged and older people: a systematic review. Front Psychol. 2022;12:797537. doi:10.3389/fpsyg.2021.797537

29. Li X, Bae JH, Lim B, Seo J, Sung Y, Jiang S, et al. Impact of Taekwondo training on cognitive and physical function in elderly individuals: a comprehensive review of randomized controlled trials. Complement Ther Clin Pract. 2024;57:101878. doi:10.1016/j.ctcp.2024.101878

30. Kumari BNP, John KKS, ManikyaLatha J, Divya PD, Maria Therese A, Arulappan J. Effect of yoga and exercises to improve physical function and quality of life in elderly: a systematic review of randomized controlled trials. Indian J Public Health Res Dev. 2022;13(3):328–331. July 31, 2025 at 10:01 AM

31. Rogers CE, Larkey LK, Keller C. A review of clinical trials of Tai Chi and Qigong in older adults. West J Nurs Res. 2009;31(2):245–279.

32. Merom D, Stanaway F, Gebel K, et al. Supporting active ageing before retirement: a systematic review and meta-analysis of workplace physical activity interventions targeting older employees. BMJ Open. 2021;11(6):e045818. doi:10.1136/bmjopen-2020-045818

33. Pucci GCMF, Neves EB, Saavedra FJF. Effect of Pilates method on physical fitness related to health in the elderly: a systematic review. Rev Bras Med Esporte. 2019;25(1):76–85. doi:10.1590/1517-869220192501193516

34. Sánchez-Sánchez JL, He L, Morales JS, de Souto Barreto P, Jiménez-Pavón D, Carbonell-Baeza A, et al. Association of physical behaviours with sarcopenia in older adults: a systematic review and meta-analysis of observational studies. Lancet Healthy Longev. 2024;5:e108–e119. doi:10.1016/S2666-7568(23)00241-6

35. Kuiper LM, Smit AP, Bizzarri D, van den Akker EB, Reinders MJT, Ghanbari M, et al. Lifestyle factors and metabolomic aging biomarkers: meta-analysis of cross-sectional and longitudinal associations in three prospective cohorts. Mech Ageing Dev. 2024;220:111958.

36. Ye KX, Sun L, Wang L, Khoo ALY, Lim KX, Lu G, et al. The role of lifestyle factors in cognitive health and dementia in oldest-old: a systematic review. Neurosci Biobehav Rev. 2023;152:105286.

37. Ibáñez A, Maito M, Botero-Rodríguez F, Fittipaldi S, Coronel C, Migeot J, et al. Healthy aging meta-analyses and scoping review of risk factors across Latin America reveal large heterogeneity and weak predictive models. Nat Aging. 2024;4:92–105.

38. Díaz Abrahan V, Bossio M, Justel N. Hacia un envejecimiento saludable: una revisión sistemática sobre la música y el ejercicio físico como factores moduladores. Actual Psicol. 2019;33(127):113–141. doi:10.15517/ap.v33i127.34975

39. Chastin S, Gardiner PA, Harvey JA, Leask CF, Jerez-Roig J, Rosenberg D, et al. Interventions for reducing sedentary behaviour in community-dwelling older adults. Cochrane Database Syst Rev. 2021;(6):CD012784. doi:10.1002/14651858.CD012784.pub2

40. Hudes R, Rich JB, Troyer AK, Yusupov I, Vandermorris S. The impact of memory-strategy training interventions on participant-reported outcomes in healthy older adults: a systematic review and meta-analysis. Psychol Aging. 2019;34(4):587–597. July 31, 2025 at 10:06 AM

41. Nguyen L, Murphy K, Andrews G. Immediate and long-term efficacy of executive functions cognitive training in older adults: a systematic review and meta-analysis. Psychol Bull. 2019;145(7):698–733.

42. Jones WE, Benge JF, Scullin MK. Preserving prospective memory in daily life: a systematic review and meta-analysis of mnemonic strategy, cognitive training, external memory aid, and combination interventions. Neuropsychology. 2021;35(1):123–40.

43. Karr JE, Areshenkoff CN, Rast P, Garcia-Barrera MA. An empirical comparison of the therapeutic benefits of physical exercise and cognitive training on the executive functions of older adults: a meta-analysis of controlled trials. Neuropsychology. 2014;28(6):829–45.

44. Lopes AL, Furtado GE, Vieira ER, da Silva DG, da Silva Neto LS, Benedetti TRB. Effects of dance interventions on cognition, depressive symptoms, and quality of life in older adults: a systematic review and meta-analysis. Brain Sci. 2023;13(6):981.

45. Vedel A, Larsen L, Aamand A. The efficacy of individual psychological interventions with non-clinical older adults: a systematic review. Eur Psychol. 2020;25(3):200–10.

46. Heaven B, Brown LJE, White M, Errington L, Mathers JC, Moffatt S. Supporting well-being in retirement through meaningful social roles: systematic review of intervention studies. Milbank Q. 2013;91(2):222–87.

47. Koh WQ, Wora N, Liong NWL, Ludlow K, Pachana NA, Liddle J. Non–exercise-based interventions to support healthy aging in older adults: a systematic review and meta-analysis of randomized controlled trials. Gerontologist. 2025;65(2):gnae156.

48. Lima KC, Caldas CP, Veras RP, Correa RF, Bonfada D, Souza DLB, et al. Health promotion and education: a study of the effectiveness of programs focusing on the aging process. Int J Health Serv. 2017;47(3):550–70.

49. Núñez de Arenas-Arroyo S, Cavero-Redondo I, Alvarez-Bueno C, Sequí-Domínguez I, Reina-Gutiérrez S, Martínez-Vizcaíno V. Effect of eHealth to increase physical activity in healthy adults over 55 years: a systematic review and meta-analysis. Scand J Med Sci Sports. 2021;31:776–89. doi:10.1111/sms.13903.

50. Stockwell S, Schofield P, Fisher A, Firth J, Jackson SE, Stubbs B, et al. Digital behavior change interventions to promote physical activity and/or reduce sedentary behavior in older adults: a systematic review and meta-analysis. Exp Gerontol. 2019;120:68–87.

51. Alley SJ, Waters KM, Parker F, Peiris DL, Fien S, Rebar AL, et al. The effectiveness of digital physical activity interventions in older adults: a systematic umbrella review and meta-meta-analysis. Int J Behav Nutr Phys Act. 2024;21:144. doi:10.1186/s12966-024-01694-4.

52. Toril P, Reales JM, Ballesteros S. Video game training enhances cognition of older adults: a meta-analytic study. Psychol Aging. 2014;29(3):706–16.

53. Stara V, Santini S, Kropf J, D’Amen B. Digital health coaching programs among older employees in transition to retirement: systematic literature review. J Med Internet Res. 2020;22(9):e17809.

54. Bacanoiu MV, Danoiu M. New strategies to improve the quality of life for normal aging versus pathological aging. J Clin Med. 2022;11(14):1–15.

55. Lehert P, Villaseca P, Hogervorst E, Maki PM, Henderson VW. Individually modifiable risk factors to ameliorate cognitive aging: a systematic review and meta-analysis. Climacteric. 2015;18(5):678–89. doi:10.3109/13697137.2015.1078106.

56. Robert C, Erdt M, Lee J, Cao Y, Naharudin NB, Theng YL. Effectiveness of eHealth nutritional interventions for middle-aged and older adults: systematic review and meta-analysis. J Med Internet Res. 2021;23(5):e15649. doi:10.2196/15649.

57. Caristia S, De Vito M, Sarro A, Leone A, Pecere A, Zibetti A, et al. Is caloric restriction associated with better healthy aging outcomes? A systematic review and meta-analysis of randomized controlled trials. Nutrients. 2020;12(8):2290.

58. Godos J, Micek A, Currenti W, Franchi C, Poli A, Battino M, et al. Fish consumption, cognitive impairment and dementia: an updated dose-response meta-analysis of observational studies. Aging Clin Exp Res. 2024;36:171. doi:10.1007/s40520-024-02823-6.

59. Junqueira RAC, Pinto AC. Healthy longevity: a systematic review of nutrological and lifestyle aspects. Int J Nutrol. 2023;16(3):1–9. doi:10.54448/ijn23302.

60. Dai Z, Lee SY, Sharma S, Ullah S, Tan ECK, Brodaty H, et al. A systematic review of diet and medication use among centenarians and near-centenarians worldwide. GeroScience. 2024;46:6625–39. doi:10.1007/s11357-024-01247-4.

